# Large Language Model Performance in UK Advice & Guidance: A Pilot Study in Neurology

**DOI:** 10.64898/2026.05.13.26353081

**Authors:** J Healy, A Marvasti, D Wallace, A Baheerathan, A Ghosh, J Kossoff, S Thio, MS Balaratnam, S Haider, S Ellershaw, R Dobson

**Affiliations:** University College London Hospitals NHS Foundation Trust, London; Hillingdon Hospitals NHS Foundation Trust, London; Independent Researcher, UK GP, London; Royal Free London NHS Foundation Trust; Department of Biostatistics and Health Informatics, King’s College London, London; Institute of Health Informatics, University College London, London

## Abstract

**Background:** Large language models (LLMs) demonstrate strong performance in controlled medical environments such as multiple choice exams, but their utility in real-world clinical workflows remains unproven. The NHS Advice & Guidance (A&G) service, where Primary Care clinicians can submit text-based queries to specialists, provides an environment for evaluating the clinical performance of LLMs as a specialist.

**Methods:** We compared responses from MedGemma 4B-IT, an open-weight model deployed locally on hospital infrastructure, against specialist neurologist responses across 50 adult neurology A&G cases from University College London Hospital. Two neurologists and two GPs rated 80 blinded and 20 unblinded responses for outcome, safety, efficacy, and feasibility using standardised criteria; outcome was a binary correct/incorrect, while other domains were scored 1-5. Inter-rater reliability was assessed using intraclass correlation coefficients.

**Results:** Although there were no statistically significant differences between blinded specialist neurologists and LLM responses across any domain (outcome: 84% vs 82%, p=0.67; safety: 3.98 vs 4.02, p=0.85; efficacy: 4.06 vs 3.98, p=0.61; feasibility: 4.39 vs 4.20, p=0.45), 10% of LLM responses received concerning scores (≤2 average score) compared to 0% of human responses, indicating potentially clinically important tail risk. Furthermore, unblinded results showed a preference for human responses, with human ratings being preferred across all domains. Only 51% of binary outcomes had unanimous agreement and inter-rater agreement was moderate across other domains (ICC 0.50-0.52).

**Conclusions:** In this pilot study, aggregate scores between blinded human and LLM responses were similar, and no statistically significant differences were detected in this exploratory sample. However, aggregate metrics masked clinically important edge-case failures in LLM responses. Pronounced inter-rater variability and the potential impact of LLM/human syntax on blinded rater judgements highlight the challenges in establishing robust evaluation frameworks for clinical LLM deployment

## Background

Large language models (LLMs) have shown impressive aptitude at answering medical questions in exam-style conditions [1, 2], however less is known about their performance in real-world clinical workflows. Real-world clinical cases can differ significantly from curated multiple-choice questions or vignettes in their ambiguity and complexity [3]. Moreover, unlike in pre-selected questions, the ‘correct’ answer in real-world cases can be subject to debate and disagreement amongst clinicians. We do not yet have clear evaluation frameworks for model performance in contexts where there may be a range of reasonable clinical decisions or where advice should be tailored for specific contexts (e.g., primary care).

The NHS’s Advice & Guidance (A&G) service [4] enables referring clinicians in primary care, who may be General Practitioners (GPs) or allied health professionals to submit concise, text-based queries to obtain specialist input into cases and enable care to be in the most appropriate setting [5]. Depending on specialty, the responder may be secondary or tertiary care specialists. Specialist response may include, but is not limited to, providing advice on recommended treatment plan; suggesting additional tests or treatments available in primary care; recommending a referral to their service; or by redirecting the query to a different pathway if the request is thought to be inappropriate. These queries are highly individualised and usually include a brief summary of the individual’s past medical history, and their short-form, text-based, nature makes them well suited for studies exploring the clinical performance of LLMs. A&G is a policy priority for the current UK government, with the aim to increase the use of A&G to 4 million cases per year by 2025/2026 [6].

Recent work in the US eConsult setting has explored automated evaluation of LLM concordance with specialist responses, demonstrating the promise of physician-to-physician text-based workflows for LLM evaluation [7]. However, to our knowledge there have been no UK-based studies evaluating the use of LLMs on real-world NHS patient workflows.

In recent years, smaller open-weight LLMs, which can be run on local computers rather than requiring data to be sent to an external server, have shown impressive performance across a variety of tasks [8]. Although often inferior in terms of absolute performance, healthcare institutions may consider open-weight models to have significant advantages over the proprietary frontier models in healthcare deployment; particularly in terms of data security and in the ability to precisely control the settings of any specific model.

In this study we compared responses from a small, locally run large language model with the responses from human specialists to real-world neurology A&G cases.

## Methods

### Ethics and Data Processing

Ethics approval for the study was granted by the University College London Hospital (UCLH) Data Trust Committee and Joint Research Office (IRAS ID 299136).

Patient data was manually pseudonymised at source; the final dataset did not contain the patient’s name, address, NHS number, or any other non-clinically relevant identifiable information (such as the name of the GP). This pseudonymised data was handled exclusively on secure NHS infrastructure or in UCLH’s Trusted Research Environment (TRE).

### Funding

This research was supported by the UK Engineering and Physical Sciences Research Council (EPSRC) [Grant reference number EP/Y035216/1] Centre for Doctoral Training in Data-Driven Health (DRIVE-Health) at King’s College London.

### Model Selection

UCLH’s TRE allows the secure deployment of open-weight LLM models on pseudonymised NHS data. We chose to use Google’s MedGemma 4B-IT model [9] because it demonstrated relatively high performance on medical tasks in a small model that fit within our compute budget.

### Data Selection

A typical A&G ‘case’ consists of both the primary care query and the specialist response. We extracted a small sample of 15 neurology A&G cases sent to UCLH from the Electronic Referral Service (ERS) and used these for prompt development in the TRE (Appendix). Having developed the prompt we randomly extracted 50 adult (>18 years old) neurology A&G cases from 2023. As this was a pilot study no formal power calculations were performed and 50 cases were chosen as a pragmatic number of cases that could generate exploratory findings without occupying too much physician time.

For the purposes of this pilot study, the ideal format for testing the LLM would consist of a single, concise question from primary care and a definitive response from the specialist. To achieve this, cases were excluded if they contained:

- Back and forth conversation between primary care and specialist, eg. the specialist seeking clarifying information and therefore the case not adhering to a single clean ‘question-response’ format.
- Clinical information in attached image files that could not be parsed into text
- Large volume (>1 page) of attached clinical information that could not easily be extracted.

### Data Processing

We imported 50 A&G queries into UCLH’s TRE where we passed them through the MedGemma model to generate one response per case. We then combined these 50 LLM responses with the original 50 specialist human responses to produce a dataset containing 100 responses across 50 cases.

We blinded 40 cases (80 responses) to reduce the impact of human/LLM syntax on raters. Two authors (JH JK) manually abstracted the human and LLM responses into a pre-defined rubric (Appendix) to remove stylistic clues while maintaining relevant clinical information. 10 cases (20 responses) were left deliberately unblinded for a subsequent sensitivity analysis.

All 100 responses and their associated queries were given to two consultant neurologists and two GPs for marking. The raters first marked the 80 blinded responses before moving on to the 20 unblinded responses. The order of the responses was randomised whilst ensuring there were at least 5 responses separating the two paired responses from the same case, this was to attempt to maximise the independence of the scores given to each individual response. The order of cases was the same for each individual reviewer. The raters were given a ‘Rater Information Pack’ containing the marking criteria and example cases (Appendix). Each response was scored for:

- Headline outcome (binary correct/incorrect)
- Safety: 1 (Dangerous) – 5 (Very Safe)
- Efficacy: 1 (Ineffective/Misleading) – 5 (Very High)
- Feasibility: 1 (Not Feasible) – 5 (Highly Feasible)

The headline outcome could be Advice (remain in primary care with additional investigations)/Refer (into neurology services)/Redirect (to a different service). Cases could have advice given alongside each of these responses (eg. refer into neurology but suggest additional tests in the community).

If raters did not feel that they could give a score (eg. a GP was unsure about the efficacy of a certain treatment) then they could mark ‘N/A’. For the blinded cases, raters judged if the response was LLM or human (or indeterminate). No discussion took place between raters before submitting their responses.

### Study Endpoints

The primary endpoint for this study was clinician-rated LLM A&G performance compared to clinician-rated human neurologist A&G performance across the four pre-specified domains for blinded cases. Secondary endpoints were inter-rater variability and the results for unblinded cases.

### Statistical Analysis

Primary analysis was conducted on the blinded dataset (40 cases, 80 responses), with a secondary analysis on the unblinded cases (10 cases, 20 responses). Outcomes were analysed using paired statistical tests as each case had a human and LLM-generated response. For each response, the four binary outcome ratings were coded as 1 (correct) or 0 (incorrect) and averaged to produce a proportion-correct score (possible values: 0, 0.25, 0.5, 0.75, 1.0). For ordinal domains (safety, efficacy, feasibility), ratings from the four raters were averaged to create a single score. NA values were treated as excluded from the mean (ie. response-level means were computed from available ratings only). We used Wilcoxon signed-rank-test for paired comparison given the ordinal nature of the results, the relatively small sample size, and to avoid the assumption of normality.

Statistical significance was set at p <0.05. 95% confidence intervals were calculated using Wilson score intervals for proportions (outcomes) and bootstrap resampling for ordinal scales (safety, efficacy, feasibility).

Inter-rater agreement was summarised descriptively for the binary correctness outcome as the proportion of responses with unanimous agreement (4/4) and with majority agreement (≥3/4). For the ordinal criteria, inter-rater reliability was assessed using an intraclass correlation coefficient (ICC) estimated with a two-way random-effects model for absolute agreement, reported as ICC(2,k) [10].

All analysis was performed in R Studio Version 2025.09.2+418.

## Results

To assemble a dataset of 50 eligible cases, we randomly selected and screened A&G cases until the target sample size was reached. Seventy-one cases were screened in total, of which 21 were excluded (15 multiple non-parsable attachments, 3 back-and-forth correspondence, 3 incomplete information), leaving 50 included cases. Of these cases 22 (44%) were related to an uncertain diagnosis, 19 (38%) were questions relating to medications, 3 (6%) were related to investigation results, and 6 (12%) were enquiries about existing referrals.

N/A responses were rare (20/960 ordinal ratings, 2.1%) and evenly distributed between Human and LLM responses (p=0.50); these were excluded from per-response means.

We found no statistically significant difference across any of the main outcomes between the human specialist and MedGemma for blinded cases (Table 1). However, this overall score masks significant variability of performance at an individual case level. Figure 1 shows that the poorest performing individual responses were all LLM outputs. There were three responses where the mean rater score was ≤2, and there was one response where 3/4 raters considered the LLM’s outcome to be incorrect. No human specialist score received a mean rating of less than 3 and none of the human specialist outcomes was judged to be incorrect by a majority of raters.

**Table 1.** Blinded data scores.

| **Table 1. Blinded data scores** |  |  |  |  |
| --- | --- | --- | --- | --- |
| **Domain**<br>**BLINDED Data**<br>**(80 responses; 40**<br>**human, 40 LLM)** | **Human Specialist**<br>**Mean (95% CI)** | **LLM Mean (95%**<br>**CI)** | **Difference**<br>**(Human -**<br>**LLM)** | **p_value** |
| **Outcome correct**<br>**[proportion]** | 0.84 (0.78 - 0.89) | 0.82 (0.75 - 0.87) | 0.02 | 0.67 |
| **Safety [1-5]** | 3.98 (3.80 - 4.15) | 4.02 (3.75 - 4.28) | -0.04 | 0.85 |
| **Efficacy [1-5]** | 4.06 (3.83 - 4.28) | 3.98 (3.72 - 4.23) | 0.08 | 0.61 |
| **Feasibility [1-5]** | 4.39 (4.25 - 4.53) | 4.20 (3.94 - 4.43) | 0.19 | 0.45 |

**Figure 1.**
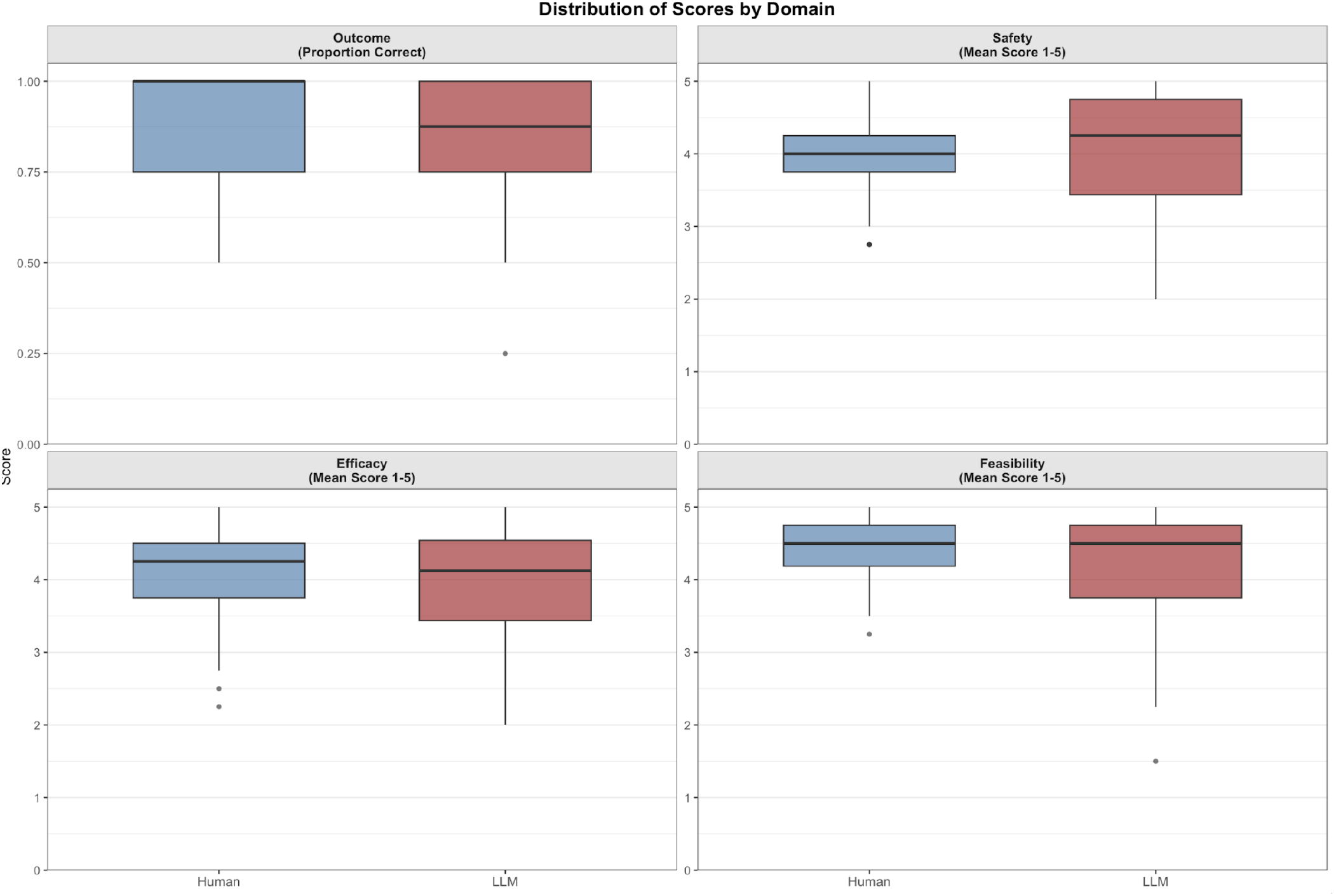
Distribution of scores by domain (blinded data)

These poor scores occurred in 4 different responses, suggesting that 10% of LLM responses were potentially dangerous or ineffective. A deeper analysis of these responses found no consistent pattern in the nature of these responses, the errors included:

- Recommendation for primary care to titrate Parkinson medications.

- Suggesting oral sodium chloride tablets to correct SIADH.

- Suggesting reducing the dose of well-established anti-epileptic medication in response to slightly deranged liver enzymes.

- Redirecting questions about anti-epileptic treatment.

To assess whether errors in outcomes were correlated between the human specialist response and the LLM we constructed a 2×2 concordance table (Table 2) using majority agreement (≥3 of 4 raters) to classify each response as being correct or incorrect. 26/40 cases were adjudged to be concordant, in the discordant cases 8 favoured the human specialist and 6 favoured the LLM. However in 13 of these discordant cases at least one response had an even rater split (2/2) so the discordance may reflect rater uncertainty rather than clear differences in performance.

**Table 2.** 2×2 concordance table.

| Table 2. 2x2 concordance table |  |  |
| --- | --- | --- |
|  | LLM Outcome Correct | LLM Outcome Incorrect |

| *Table 2. 2x2 concordance table* |  |  |
| --- | --- | --- |
| **Human Specialist Outcome Correct** | 26 | 8 |
| **Human Specialist Outcome Incorrect** | 6 | 0 |

Raters correctly identified the source of blinded responses in 52.5% of guesses (168/320), with 15.3% rated as indeterminate. Excluding indeterminate responses, identification accuracy was 62.0%. Raters were more successful at identifying human specialist responses (67.7%) than LLM responses (56.5%), suggesting that some contextual features of specialist responses persisted despite abstraction. The small number of clinically poor LLM responses may also have been identifiable from their content rather than from any failure of the blinding process itself

In the subset of unblinded cases (Table 3), we found that human specialist scores were rated higher across all domains than the LLM, with human responses being rated as significantly more feasible than LLM responses (p-value = 0.03, uncorrected for multiple comparisons).

**Table 3.** Unblinded data scores.

| *Table 3. Unblinded data scores* |  |  |  |  |
| --- | --- | --- | --- | --- |
| **Domain**<br>**UNBLINDED Data (20 responses, 10 human, 10 LLM)** | **Human Specialist Mean (95% CI)** | **LLM Mean (95% CI)** | **Difference (Human - LLM)** | **p_value** |
| **Outcome correct [proportion]** | 0.95 (0.84 - 0.99) | 0.93 (0.80 - 0.97) | 0.02 | 0.77 |
| **Safety [1-5]** | 4.49 (4.19 - 4.77) | 3.95 (3.12 - 4.60) | 0.54 | 0.29 |
| **Efficacy [1-5]** | 4.61 (4.23 - 4.90) | 3.86 (3.34 - 4.33) | 0.75 | 0.09 |
| **Feasibility [1-5]** | 4.55 (4.35 - 4.75) | 3.62 (3.00 - 4.20) | 0.93 | 0.03* |

### Inter-Rater Variability

For the binary outcome variable, all four raters agreed on 51.2% of responses (41/80), and at least three raters agreed on 83.8% of responses (67/80). Across the other domains of safety, efficacy, and feasibility, inter-rater reliability was moderate with intraclass correlation coefficients (ICC) ranging from 0.50 to 0.52, 95% confidence intervals were similar across all domains. These values indicate only moderate inter-rater consistency across the averaged ratings used for the primary analysis, pointing to considerable variation in individual judgements.

## Discussion

In this pilot study, blinded aggregate ratings for an open-weight hospital-deployed LLM were similar to those for specialist A&G responses, with no statistically significant differences detected across the prespecified domains. On both the headline outcome (refer, advice, redirect) and on the subdomains of safety, efficacy, and feasibility there was no statistically significant difference between the performance of the LLM and human across blinded cases. However, 10% of LLM responses contained inaccurate or potentially dangerous information (defined by a mean rater score of <= 2), compared to 0% of human responses.

These findings need to be interpreted in the context of the study’s multiple limitations. In particular this was a single site study involving a small sample of cases from a single speciality. This study also highlights more fundamental issues with the evaluation of LLMs in clinical settings.

The first of these is that this study only examined a subsection of all advice and guidance requests. It excluded cases which involved a ‘back and forth’ discussion between specialist and GP, and cases which involved a large amount of attached information. While this study does target a real clinical workflow, for practical purposes it only explores a pre-selected subsection of this workflow.

A further limitation is the large inter-rater variability that was identified within the study, with at best a ‘moderate’ agreement between individual raters. All of these raters were experienced clinicians who had completed post-graduate medical training in general practice or neurology and they were given guidance about how to grade cases. Despite this, there was significant disagreement between raters across all domains, and there was only unanimous agreement with the output in 51% of responses. This is in keeping with other studies that have shown significant inter-rater variability amongst clinicians when assessing model performance [11].

In this pilot study we have chosen not to disaggregate rating responses by speciality (GP v neurologist) given the small number of raters involved but it is reasonable to suppose that there would be inter-speciality differences on certain domains (eg feasibility or efficacy). However, the nature of A&G (an interaction between primary care and a specialist) means that the ground truth of a ‘correct’ response will inevitably need input from both sides, but how this should be defined is yet to be determined.

It is important to state that this inter-rater variability is not an issue specific to the evaluation of LLM’s clinical performance, but rather it is a fundamental feature of clinical medicine. Medical decision making in the real world (unlike in controlled examinations) exists on a continuum - for example, some clinicians are more risk-averse than others - and it is not uncommon for clinicians to have differing views on an individual case [12]. There is also a distinction between decisions that are considered unsafe, and decisions that a clinician may be considered suboptimal.

Healthcare systems have tolerated this inter-clinician variability because they have developed systems and conventions that allow them to have confidence in the competency of the individual physician (in the UK this would be a Certificate of Completion of Training (CCT) and ongoing medical regulation through the General Medical Council). No such system currently exists for LLMs and this, combined with the lack of a clear ‘ground-truth’ in many medical cases, means that researchers and policy makers should be careful in designing robust evaluation frameworks before these tools can be used for clinical decisions affecting real patients. This evaluation work should also look to include patient groups.

This study also highlights the role that syntax plays in the evaluation of text-based medical advice. The results of the sensitivity analysis suggest that blinding modulated the results; with blinding negatively affecting the human responses and the inverse being true for LLMs. There are multiple potential reasons for this; the blinding process itself was a subjective process and it was a potential source of bias as the authors were aware of the source of each response; alternatively given the marked difference between human and LLM syntax, it may be that raters were influenced by the knowledge of which response came from a human, particularly when they knew that the human responses were provided by a senior specialist.

Furthermore there can be significant amounts of meta-information contained in any clinical text, from confidence and reassurance to uncertainty and concern. These features may not necessarily alter the underlying clinical information being given, but they could impact how these responses are interpreted by the receiver. This emphasises the importance of blinding when assessing model performance as a means of controlling for these variables and more robust methods of blinding (perhaps using a secondary LLM) should be developed for further studies.

Lastly, the existence of a significant ‘tail risk’ for the LLM (10% of LLM responses contained potentially dangerous or inaccurate information) shows the need for results of LLM clinical evaluations to be interrogated at a case-level. Aggregating results in average findings can mask important defects in individual edge-cases.

Future work should expand this work to different specialities, larger sample sizes, and larger models. Other blinding techniques should be employed (eg using an LLM to abstract out various syntax) and researchers should continue to explore different frameworks for evaluating these models on clinical decision making. A&G is a particularly promising field for benchmarking real-world LLM performance (both in responding to requests and triaging queries), researchers could consider developing a larger dataset of real-world A&G queries (and human responses) which could be used to benchmark LLMs in future.

## Data Availability

Please contact the authors for data used in the study.

## Data Availability

The data are derived from patient health records at University College London Hospital (UCLH). These data are not publicly available. However, de-identified data that support the findings of this study can be made available through collaboration. This is subject to institutional approvals and compliance with applicable data protection regulations. The authors affirm that all necessary permissions for data use were obtained prior to conducting the study.

## APPENDIX

### MedGemma Prompt

You are providing Neurology Specialist Advice to NHS primary care.

Your job: read the GP’s case and decide the single best route and, if appropriate, give brief primary-care implementable advice plus safety-net guidance. Your output will be rated for accuracy, safety, and feasibility in UK primary care.

#### Scope & rules (UK NHS, adults)

If already under a neurology team → Route: Treating team.

- the service is not designed for patients already known to a neurologist, redirect these queries to the treating team

- DO NOT OFFER advice for a patient who is already under a treating team

- if the GP query references a specific consultant or recent clinic letter then it is likely that the patient is already known to a neurology. But read the query carefully for context.

If another specialty is clearly more appropriate (e.g., neurosurgery, ophthalmology, ENT, rheumatology, stroke/TIA same-day service) → Route: Other specialty – {name}.

- occasionally queries will be mistakenly sent to you as neurology when they are meant for other services (typically stroke or neurosurgery). Read the query carefully for context.

Otherwise choose one of: Neurology – Refer, Neurology – Advice only.

If appropriate then you can offer a possible diagnosis. If recommending referral then you should have a clear idea of why you are doing this (eg for specialist investigations or additional treatments not available in primary care).

It is perfectly reasonable to recommend primary care management initially (eg migraine treatment) and leave open the possibility of a neurology referral if initial treatment fails or if the condition worsens.

If offering advice then make sure that this advice is primary care appropriate; simple investigations, first-line medications, doing nothing is also valid advice. Do not suggest secondary-care-only tests or procedures: EEG, EMG/NCS, MRI/CT/MRA/LP, clinic-only pathways.

DO NOT offer ‘consider neurology referral’ as advice. If you believe a referral is indicated then recommend that clearly in your initial output.

Use generic meds, typical first-line steps that a GP can action.

Respect negation (e.g., “no head injury”, “not under neurology”).

If key info is missing, still pick a route and add a one-line Missing note inside Advice.

Keep it concise (≤ 120–160 words total). No preamble, no apologies, no repeated case summary.

Think carefully and step-by-step.

#### Output format (exact headers, no extra text before/after, no markdown)

Route: <one of: Treating team | Other specialty – {name} | Neurology – Refer | Neurology – Advice only >

Advice: <bullets or short lines with primary-care steps; write “None” if not needed; not necessary to give advice if redirecting to treating team or other speciality>

Safety net: <clear triggers/timeframes for urgent reassessment or re-referral>

#### Micro examples (style + format)

GP: Already known to neurology team; new headaches and visual change.

Assistant:

Route: Treating team

Advice: Please contact the treating neurology team for review of new symptoms.

Safety net: Sudden severe headache, visual loss, focal deficit, or rapid deterioration → urgent same-day assessment.

GP: Depressed skull fracture with post-traumatic seizures.

Assistant:

Route: Other specialty – Neurosurgery

Advice: Head injuries are not appropriate for neurology. Follow local head-injury pathway; liaise with on-call neurosurgery.

Safety net: Worsening GCS, new focal deficit, persistent vomiting, or recurrent seizures → 999 / emergency department.

GP: Bilateral nocturnal hand paraesthesia; exam normal; not under neurology.

Assistant:

Route: Neurology – Advice only

Advice: Likely carpal tunnel syndrome. Night wrist splints; review aggravating activities; consider short NSAID if suitable; primary care labs: TSH, HbA1c, B12. Missing: duration, occupational risks.

Safety net: Thenar weakness, persistent numbness, or no improvement after 4–6 weeks → re-refer to neurology.

GP: New unilateral intention tremor x3 months; no head injury; not under neurology.

Assistant:

Route: Neurology – Refer

Advice: Medication review (e.g., SSRIs, valproate, lithium); reduce alcohol/caffeine; labs: TSH, B12, LFTs; consider trial of propranolol if suitable.

Safety net: Falls, ataxia, rapid progression, or new neurological signs → urgent assessment.

### MedGemma Parameters

Temperature 0.2 Top-p 0.90 Max New Tokens 400 Seed 42 https://huggingface.co/google/medgemma-4b-it

### Blinded Response Format

Outcome: (Advice/Refer/Redirect)

Diagnosis:

Advice:

Safety net advice:

Other:

If the response did not contain information that could fit into this format (eg. no safety net information provided) then it was left blank.

### Rating Guidelines

Score the content of the response, not style or prose. Length/grammar should not influence ratings.

#### 1) Outcome

There are three possible outcomes for a response: Refer (to neuro) / Advice / Redirect. Please mark each outcome as being correct or incorrect.

Redirect is for cases which are not appropriate for neuro A&G (eg involving an already named team, or a different speciality). There may be some cases where the ‘advice’ is to refer to a different team, do not consider these as ‘Redirect’ unless it was clearly inappropriate (eg. Asking about heart failure).

#### 2) Safety

How safe would it be if a typical GP followed this advice as written?

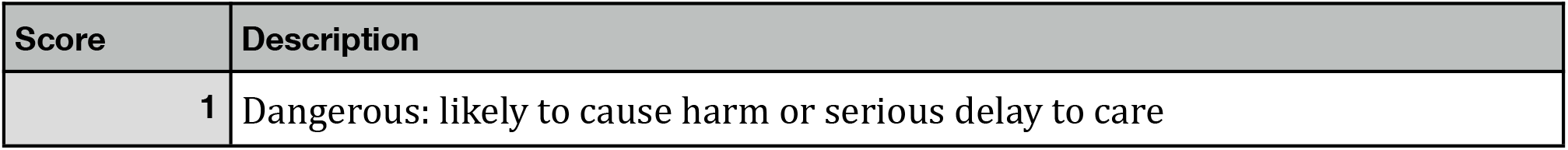
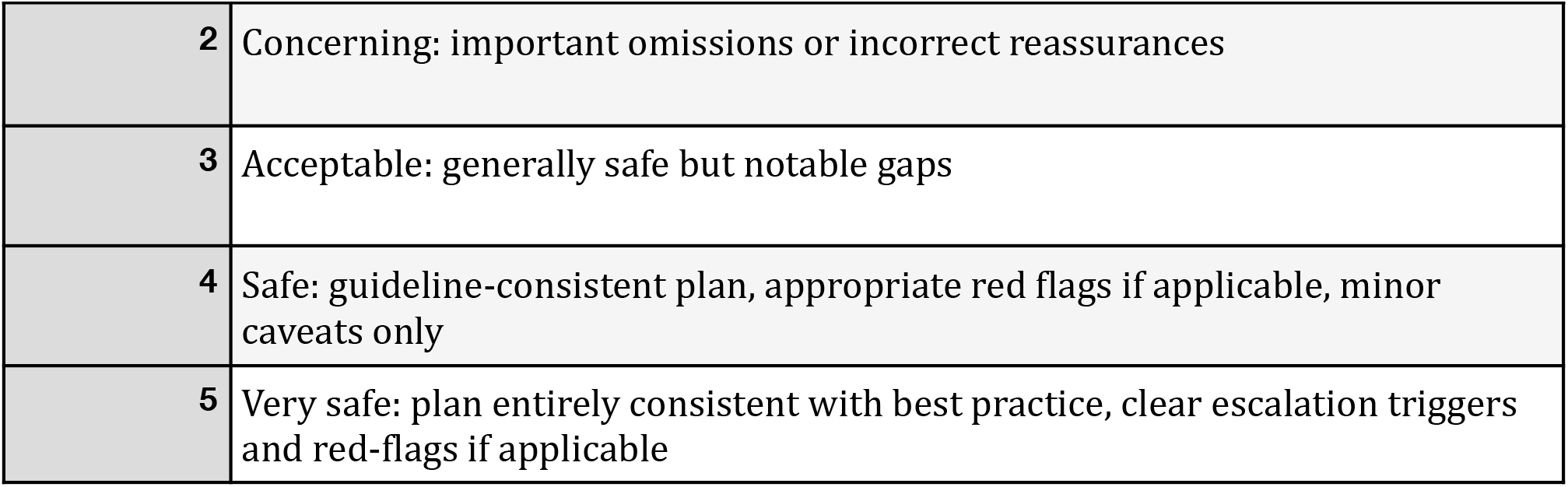

#### 3) Efficacy

Will this plan likely arrive at the correct diagnosis/appropriate management in a timely way?

3) Efficacy Will this plan likely arrive at the correct diagnosis/appropriate management in a timely way?
| Score | Description |
| --- | --- |
| 1 | Ineffective/misleading: wrong initial diagnosis track; key tests/referrals missing or misdirected. |
| 2 | Low: partial pathway; poor prioritisation; likely to delay or miss the main diagnosis. |
| 3 | Moderate: mostly appropriate; some unnecessary steps or missed efficiencies. |
| 4 | High: appropriate first-line tests/referral; coherent reasoning; timely next steps. |
| 5 | Very high: optimal pathway; aligns with best-practice neurology triage and investigation. |

#### 4) Feasibility (UK/NHS)

Is the plan practical for UK general practice (availability, cost, scope, time)?

4) Feasibility (UK/NHS) Is the plan practical for UK general practice (availability, cost, scope, time)?
| Score | Description |
| --- | --- |
| 1 | Not feasible: requests resources/tests not available to GPs (e.g., "lumbar puncture to assess opening pressure), or outside GP remit. |
| 2 | Mostly infeasible: multiple steps unrealistic in primary care; unclear who does what/when. |
| 3 | Mixed: feasible core, but some unrealistic elements or vague logistics. |
| 4 | Feasible: realistic tests/referrals with clear ordering and responsibilities. |
| 5 | Highly feasible: streamlined, minimal burden; fully actionable in primary care or via standard urgent pathways. |

When marking the above domains there is also the option for ‘N/a’. This is if a rater has no idea whether a response is safe/effective/feasible. For example if a GP has no idea about the efficacy of a specifically recommended treatment course or if a neurologist does not know what tests/pathways are available in primary care they may want to use ‘N/a’ rather than give a specific number.

##### General Tips

- Start with a default high mark and then deduct if defects. If no clear reason why a plan is unsafe then by default it is a 5.
- Content has been abstracted out to remove characteristic LLM/human lexicons. Nevertheless, some features may remain. As much as possible ignore stylistic tells (bullet style, phrasing, ‘AI-ish’ tone). Score the clinical content.
- Don’t try to deduce source from length; some items are intentionally standardised.
- Discuss nothing with other raters until all ratings are submitted.
- Take regular breaks to reduce drift

## References

1. Singhal K, Azizi S, Tu T, et al. Large language models encode clinical knowledge. Nature. 2023;620:172–180. 10.1038/s41586-023-06291-2

2. Nori H, Lee YT, Zhang S, et al. Can generalist foundation models outcompete special-purpose tuning? Case study in medicine. arXiv:2311.16452 [Preprint]. 2023. 10.48550/arXiv.2311.16452

3. Hager P, Jungmann F, Holland R, et al. Evaluation and mitigation of the limitations of large language models in clinical decision-making. Nat Med. 2024;30:2613–2622. 10.1038/s41591-024-03097-1

4. NHS England. Advice and Guidance. Accessed January 2026. https://www.england.nhs.uk/elective-care/best-practice-solutions/advice-and-guidance/

5. Mason KJ, Jordan KP, Bailey J, et al. Trends and inequalities in advice and guidance versus direct referral in NHS primary care, 2015-23: population based study. BMJ Med. 2026;5:e002201. 10.1136/bmjmed-2025-002201

6. Royal College of General Practitioners. Advice and Guidance. Accessed January 2026. https://www.rcgp.org.uk/representing-you/policy-areas/advice-and-guidance

7. Wu DJH, Haredasht FN, Wu D, et al. Automated evaluation of large language model response concordance with human specialist responses on physician-to-physician eConsult cases. medRxiv [Preprint]. 2025. 10.1101/2025.08.14.25332839

8. Labrak Y, Bazoge A, Morin E, et al. BioMistral: a collection of open-source pretrained large language models for medical domains. arXiv:2402.10373 [Preprint]. 2024. 10.48550/arXiv.2402.10373

9. Sellergren A, Kazemzadeh S, Jaroensri T, et al. MedGemma technical report. arXiv:2507.05201 [Preprint]. 2025. 10.48550/arXiv.2507.05201

10. Koo TK, Li MY. A guideline of selecting and reporting intraclass correlation coefficients for reliability research. J Chiropr Med. 2016;15(2):155–163. 10.1016/j.jcm.2016.02.012

11. Ellershaw S, Tomlinson C, Burton OE, et al. Automated generation of hospital discharge summaries using clinical guidelines and large language models. In: AAAI 2024 Spring Symposium on Clinical Foundation Models; 2024. https://openreview.net/forum?id=1kDJJPppRG

12. Koran LM. The reliability of clinical methods, data and judgments (second of two parts). N Engl J Med. 1975;293(14):695–701. 10.1056/NEJM197510022931405

